# Single-cell dissection of the immune response after acute myocardial infarction

**DOI:** 10.1101/2023.05.02.23289370

**Authors:** Irene V. van Blokland, Roy Oelen, Hilde E. Groot, Jan Walter Benjamins, Kami Pekayvaz, Corinna Losert, Viktoria Knottenberg, Matthias Heinig, Leo Nicolai, Konstantin Stark, Pim van der Harst, Lude H. Franke, Monique G. P. van der Wijst

## Abstract

The role of the immune system during and in response to acute myocardial infarction (MI) is poorly characterized but is an important driver of recurrent cardiovascular events. Anti-inflammatory drugs have shown promising effects on lowering this recurrency risk, but broadly impair the immune system and may induce severe side effects. To overcome these challenges a more detailed understanding of the immune response to myocardial infarction is needed.

For this, we compared peripheral blood mononuclear cell (PBMC) single-cell RNA-sequencing expression and plasma protein profiles over time (hospital admission, 24h and 6-8 weeks after STEMI) in 38 patients and in comparison to 38 controls (95,995 diseased and 33,878 control PBMCs). Compared to controls, we observed a relative increase in classical monocytes and a decrease in CD56^dim^ natural killer cells in STEMI patients at admission, and these differences persisted until 24h after STEMI. The monocytes also showed the largest gene expression changes in each of the conditions, which was associated with changes in toll-like receptor, IFN and IL-1 signaling activity. Finally, a targeted protein cardiovascular biomarker panel revealed 33/92 plasma proteins to be changed after STEMI. Interestingly, three of these proteins were found to be affected by coronary artery disease-associated genetic risk variation, disease status and time after STEMI. Indicating the importance of taking all these aspects into consideration when defining potential future therapies.

Altogether, our analyses have revealed the immunological pathways that are disturbed upon MI, and in which cell type and during which stage of the disease this occurs. Additionally, we also provide insights in which patients are expected to benefit most from anti-inflammatory treatments, by identifying the genetic variants and disease stage at which these variants affect the outcome of these (drug-targeted) pathways. These findings advance our knowledge of the immune response after MI and provide further guidance for future therapeutic studies.

## Introduction

Acute myocardial infarction (MI) is one of the leading causes of mortality and morbidity globally. The immune system plays an important role in the pathophysiology of the disease: from the initial formation and rupture of atherosclerotic plaques, all the way up to inflammation in response to the MI (1–3). Systemically studying and targeting the inflammatory response has remained challenging due to its complexity and severe side effects, as was seen in the CANTOS trial (interleukin-1ß, IL-1ß) and ASSAIL-MI (interleukin 6 receptor, IL6R) trial (4,5). Identifying specific immune cell subpopulations and underlying processes that are specifically disturbed after MI could open the way for more targeted treatment that is expected to cause fewer side effects (6,7).

However, until recently, analyzing the characteristics of specific cell types was often done at bulk level and using predefined marker genes, not providing an unbiased, comprehensive overview of the immune system (8). Recent developments in the field of single-cell RNA sequencing (scRNA-seq) now enable us to unbiasedly study 100,000s of individual cells simultaneously on a transcriptome-wide level (9–11). This technology enables us to gain new insights into the inflammatory response after a MI by flexibly and concurrently mapping MI-induced changes in cell type composition, gene expression level and downstream pathways at various cellular resolutions.

To study MI at the single-cell level, it is essential to first understand the normal physiological variation among the general population. Such groundwork has been generated both for the heart and the circulating immune cells (9,13). Various aspects in the pathophysiology of myocardial infarction have now been dissected at the single-cell level, from atherosclerotic aortas precluding the disease (14,15) to cardiac neovascularization by resident endothelial cells after the infarction (16). More recently, single-cell multi-omic maps (combining spatial transcriptomics, single-cell expression and chromatin accessibility profiling) have been generated from human heart tissue after MI (17). This revealed an increased spatial dependency between lymphoid and myeloid cells in ischaemic samples compared to healthy samples, indicating the importance of cellular communication between immune cells during cardiac repair after MI. Altogether, these previous studies have mainly focused on the heart, whereas each of these studies also indicate the importance of the immune cells during a MI. However, we still lack a more detailed single-cell level description of how the circulating immune cells are affected by a MI, both during the acute and chronic phases of the disease.

Here, we analyzed scRNA-seq data of 95,995 peripheral blood mononuclear cells (PBMCs) from 38 ST-elevated myocardial infarction (STEMI) patients (during hospital admission, 24h and 6-8 weeks after STEMI) with those of 33,878 PBMCs from 38 age- and sex-balanced general population controls. This comparison revealed large changes in cell type composition, gene and plasma protein expression levels and cell-cell communication, both in comparison to controls and during the course of the disease (**Figure S1**). Moreover, we show that several important plasma proteins during the disease are affected by a combination of genetics, disease status or phase of the disease. Altogether, the current study increases the resolution by which the immune system during and after MI has been dissected and emphasizes the importance of taking person- and disease-related characteristics into account to fully grasp the observed underlying molecular changes.

## Methods

### Study population

All patients presenting with a first STEMI upon admission at the Heart Catherization center at the University Medical Center Groningen (UMCG) were enrolled in the study between January 1st 2018 and November 30th 2019. Inclusion criteria were adults (>18 years) that had a STEMI, a primary percutaneous intervention (PCI) with implantation of at least one stent with a diameter of at least 3 mm resulting in thrombosis in myocardial infarction (TIMI) flow grade 2 or 3 post PCI, and which showed less than 6 hours symptoms before undergoing PCI. Major exclusion criteria were previous MI, medical history of diabetes, inflammatory disease or malignancies, medication affecting inflammation and clemastine or desloratadine use during intervention. Using these criteria, we initially enrolled 88 STEMI patients in this study, of which 42 were later excluded due to missing follow-up time points (t24h, t8w). Further 8 patients were excluded in the follow-up due to presence of exclusion criteria that were not yet known upon admission (**Figure 1**). Patient data was collected and managed using REDCap electronic data capture tools hosted at the UMCG (18,19). The study was part of CardioLines, a single-center observational biobank aimed to study potential factors related to success or failure of diagnosis and treatment both from a patient as well as a medical perspective (20). The study was approved by the ethics committee of the UMCG, document number METC UMCG 2012/296. Informed consent was obtained of all patients. scRNA-seq data from 38 age- and sex-balanced participants from the LifeLines DEEP cohort were included as a control group (21,22).

**Figure 1.**
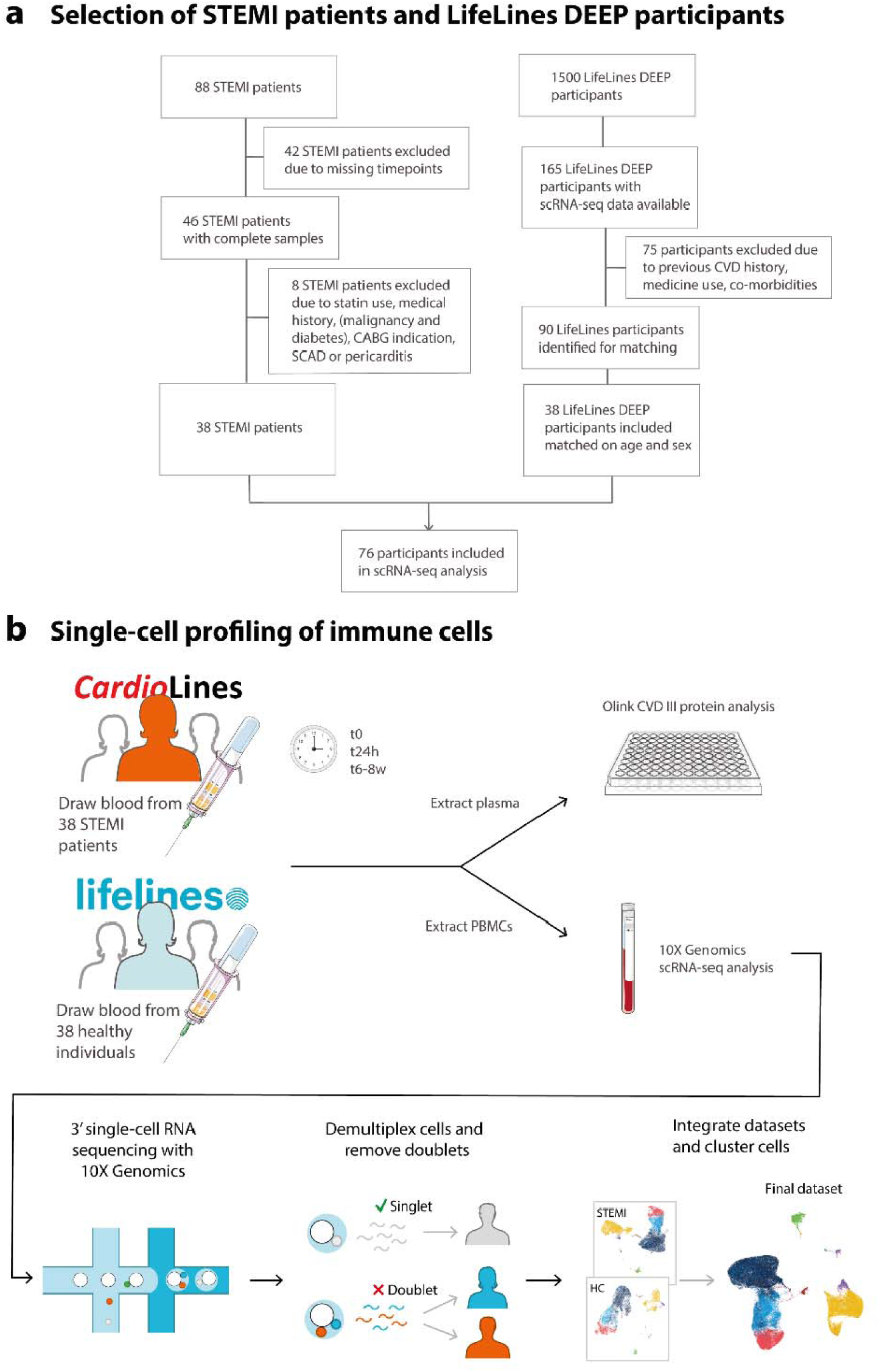
Study overview. **a.** Flow diagram for the selection of STEMI patients (left) participating in the CardioLines Biobank and controls participating in the LifeLines DEEP biobank (right). **b.** Blood was drawn from 38 STEMI patients participating in the CardioLines Biobank at three different timepoints after STEMI (hospital admission, 24 hours, 6-8 weeks post STEMI). Plasma was isolated for Olink protein analysis and PBMCs were isolated for 10X Genomics 3’-end scRNA-seq. After sequencing, samples were demultiplexed and doublets were identified. Similarly, previously Olink and scRNA-seq data was generated for the 38 age- and sex-balanced controls from the LifeLines DEEP cohort (19). Cell type classification was then performed on the QCed dataset by clustering the cells per condition in STEMI patients and controls. The cell type labels were subsequently transferred back to the dataset containing all cells.

### Clinical parameters

Upon inclusion of STEMI patients, whole blood samples were collected at three different timepoints: during admission at the heart catheterisation center (t0), 24 hours (t24h, acute phase) and 6-8 weeks (t8w, chronic phase) later (**Figure 1**). Standard laboratory assessment of the blood was performed upon admission and routine physical parameters were recorded **(Table 1)**. Titres of creatine kinase (CK), myocardial band of CK (CK-MB) and troponin T were routinely measured at 3h, 6h, 9h, 12h, 24h and 48h after admission.

**Table 1.**
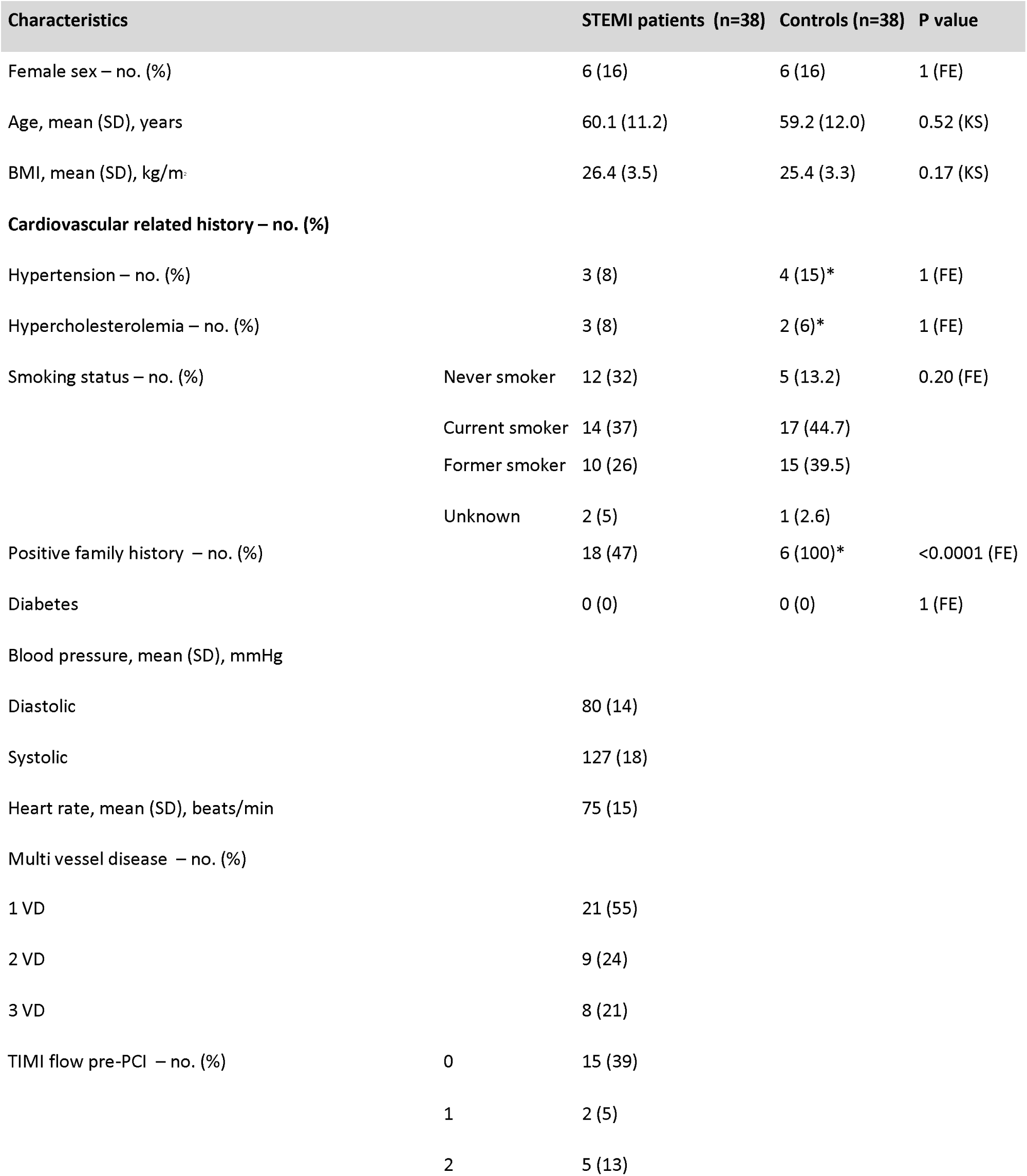
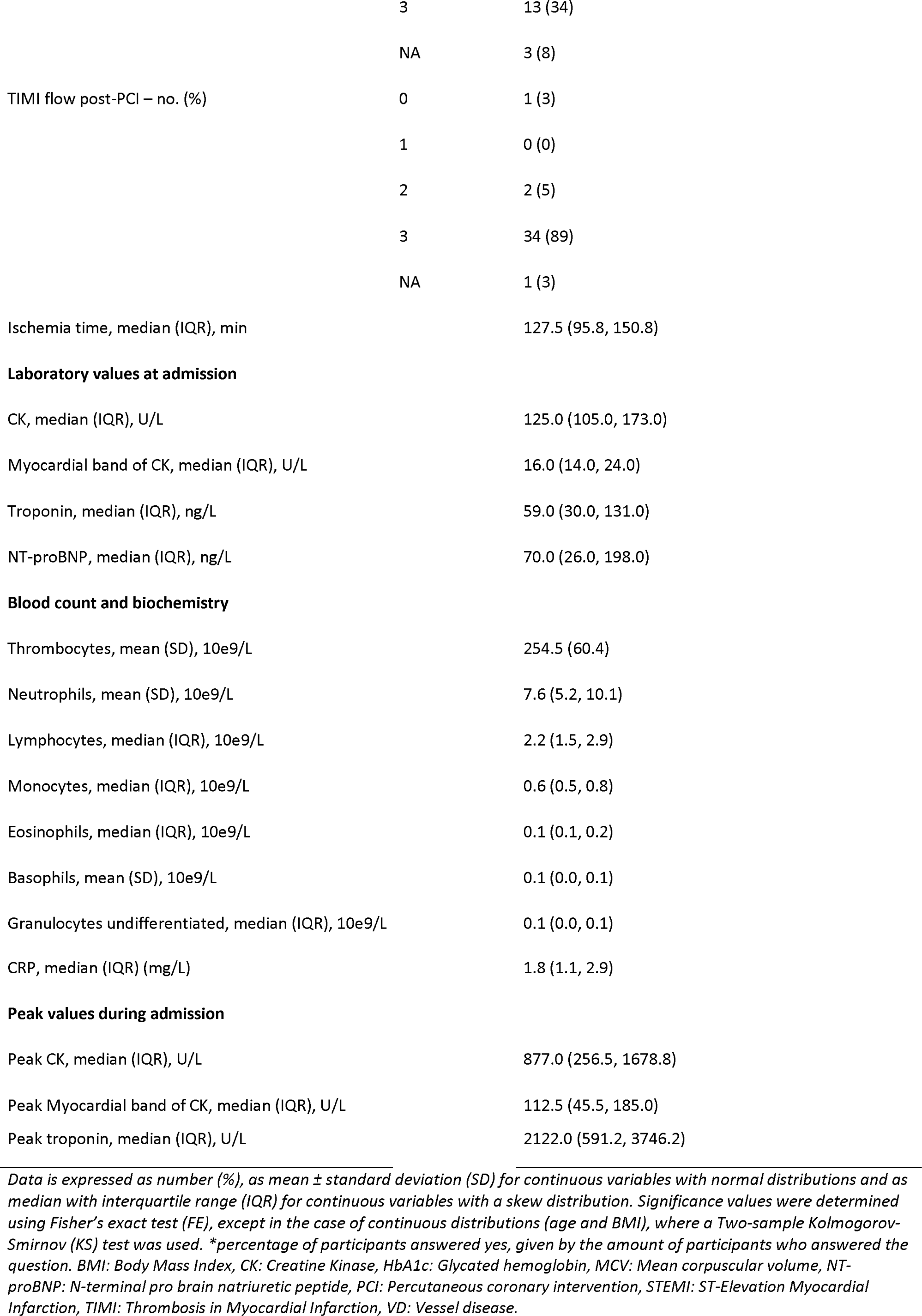
Baseline characteristics of STEMI patients and controls from the LifeLines DEEP Cohort.

### Isolation and preparation of PBMCs

PBMCs were isolated and stored as previously reported (9). In short, for each donor and timepoint, whole blood was collected in two 10 ml EDTA-vacutainer (BD) tubes. Within 2 hours after collection, PBMCs were isolated from the blood using cell preparation tubes with sodium heparin (BD). PBMCs were kept in RPMI1640 supplemented with 50⍰µg/mL gentamicin, 2 mM l-glutamine, and 1⍰mM pyruvate. Isolated PBMCs were cryopreserved in RPMI1640 containing 40% FCS and 10% DMSO. Within one year, PBMCs were further processed for scRNA-seq analysis. Cells were thawed in a 37°C water bath until almost completely thawed, after which the cells were washed with pre-warmed (37°C) RPMI1640. After washing, cells were resuspended in pre-warmed RPMI1640 medium and incubated for 1⍰h in a 45° slant rack at 37°C in a 5% CO_2_ incubator. After resting, cells were washed twice in medium supplemented with 0.04% bovine serum albumin. Cells were counted using a hemocytometer and cell viability was assessed by the Trypan Blue assay (**Figure S2**).

### Single-cell library preparation and sequencing

Fifteen sex- and timepoint-balanced sample pools were prepared aiming for 1,000 cells per patient for a total of 8 patients per pool. Single cells were captured with the 10X⍰Chromium controller (10x⍰Genomics) according to the manufacturer’s instructions (document CG00026) and as described earlier (23). Each sample pool was loaded into a different lane of a 10X⍰chip (Single Cell A Chip Kit, PN-120236 v2 and PN-1000074 v3), divided over 15 10X runs in total. cDNA libraries for v2 and v3 were generated with the Single Cell 3⍰ Library & Gel Bead kit (v2: 120237, v3: 1000075) and i7 Multiplex kit (120262) according to the company’s guidelines. These libraries were sequenced using a custom program (28-8-150 (v3) vs 27-9-150 (v2)) on an Illumina NovaSeq6000 using a 150-bp paired-end kit.

### Alignment and initial processing of sequencing data

CellRanger v3.0.2 software with default settings was used to demultiplex the sequencing data, generate FASTQ files, align the sequencing reads to the hg19 reference genome, filter cell and UMI (unique molecular identifier) barcodes, and count gene expression per cell. Genotypes of the samples were generated using the Infinium Global Screen Array-24 kit (v1 for the samples processed with 10X v2 chemistry and v3 for the samples processed with 10X v3 chemistry) and were phased using Eagle v2.4 and imputed with the HRC 1000G Phase-3 v5 (hg19) reference panel using the Michigan Imputation Server (Minimac v3) (24). Genotype information of the controls was processed as described in the Lifelines DEEP cohort paper (21). Doublet detection and sample assignment were done using Souporcell (25), confining ourselves to only use exonic single nucleotide polymorphisms (SNPs) with a MAF≥0.05. The detected doublets were discarded. Then, samples were assigned to the cell cluster with the highest correlation between the genotype retrieved from the cell cluster versus the genotypes from the individuals that had been pooled in the same 10X lane.

### Data preparation and quality control

Version 4.0 of the R package Seurat was used for data preprocessing (26–28). We performed dimensional reduction and clustering to integrate four datasets (v2 and v3 10X chemistry, both for STEMI patients and controls). For this, the Pearson residuals were used resulting from running SCTransform in each dataset using Seurat’s SCTintegration method (29). The first 30 principal components were used for cell clustering using Seurat’s FindNeighbors (dims = 1:30) and FindCluster function (default parameters, resolution 1.2) and an UMAP plot was used to visualize this. All genes that were not detected in at least 3 cells were removed. Cells with a high percentage of reads mapping to mitochondrially-encoded genes (>8% for 10X V2 and >15% for 10X V3) were discarded, as this can be a marker of poor-quality cells (30). Also, cells expressingL≤200 genes were considered outliers and discarded. Furthermore, cells expressing ≥10 UMIs of the HBB gene were discarded, indicative of erythrocytes. Finally, all cells that were marked as doublets or inconclusive by the SoupOrCell method were discarded (25). In total, 129,873 cells were used for downstream analysis consisting of 95,995 cells from STEMI patients and 33,878 cells from age- and sex-balanced controls (**Table S1**).

### Cell Type Classification

Cells were annotated in each dataset separately using Seurat’s Azimuth method by projecting them on a previously annotated multimodal CITE-seq (combined scRNA-seq and protein expression) reference dataset of 162,000 PBMCs (31). These annotations were transferred to the SCTransform integrated object. Each cluster was assigned the cell type that was the most prevalent guided Azimuth annotation in that cluster. Annotations were confirmed using marker gene expression (**Figure S3**). As a result of discordant marker gene expression, two clusters were reassigned: cluster 25 from ncMono to cMono and cluster 17 from doublets to ncMono. Major cell types were defined as “B” (B naive, B intermediate, B memory), “CD4^+^ T-cell” (Treg, CD4^+^ Naive, CD4^+^ TCM, CD4^+^ TEM, CD4^+^ CTL, CD4^+^ Proliferating), “CD8^+^ T-cell” (MAIT, CD8^+^ Naive, CD8^+^ TCM, CD8^+^ TEM, CD8^+^ Proliferating), “DC” (ASDC, cDC1, cDC2, pDC), “HSPC” (HSPC), “monocyte” (cMono: CD14^+^ and ncMono: CD16^+^), “NK” (NK dim, NK bright, NK proliferating), “plasmablast” (plasmablast), “platelet” (platelet) and “T-other” (dnT, gdT, ILC).

### Cell type abundance

Cell type proportion differences across STEMI timepoints and differences between STEMI patients and controls were determined using the permutation-based method described in Farbehi et al. (32). The number of cells of each major cell type were summed across controls or patients (separately for each STEMI timepoint). Pairwise comparisons were made across all combinations of STEMI timepoints and the controls using a W-value of 0.05. Resulting p-values were Holm-Bonferroni corrected, with a corrected p-value < 0.05 being considered statistically significant.

### Differential gene expression and pathway analysis

For differential gene expression (DE) analysis library-size normalization was performed by scaling the total UMI gene expression of each cell barcode to 10,000, after which the gene expression data was log-transformed (LogNormalize in Seurat). For each cell type, pairwise DE analysis was performed using MAST comparing everything to t0 (controls, 24 hours and 6-8 weeks) (33). Due to strong technical differences in gene expression profiles between the samples that were prepared with the 10X V2 or 10X V3 protocol, DE analysis was performed separately for the samples belonging to either version of the protocol. A meta-analysis, considering only genes tested in both datasets, was performed across the V2 and V3 results using the Fisher’s Combined Probability Test. To correct for multiple testing, Bonferroni correction was applied and adjusted p-values <0.05 were considered statistically significant. The up- and downregulated DE genes were used for separate pathway analyses using the ToppFun functional enrichment tool, selecting for the REACTOME database to identify relevant pathways (34).

### Cell-to-cell communication analysis

We predicted potential cell-to-cell interactions using NicheNet analysis v1.1.0 (35). Using a background set of DE genes, NicheNet predicts potential ligand-receptor interactions and predicts whether specific cell-cell interactions led to differences in downstream gene-expression. We modified NicheNet’s FindMarkers wrapper to use the MAST implementation in Seurat in order to determine DE ligands and receptors, as well as genes downstream of cells potentially interacting with the ligands. Differential ligand and downstream expression changes were confined to genes that were present in at least 10% of the cells. The absolute log fold change between the conditions needed to be at least 0.1 to be considered for testing. We assessed potential ligand-receptor interactions between pairs found in the Omnipath database (36). We then used NicheNet to assess ligand-receptor activity, by filtering predicted ligand-receptor interactions for those ligands that were shown to have an effect on downstream gene expression.

### Protein Olink analysis

Plasma from the 38 STEMI patients was collected at each of the three timepoints (t0, t24h, t8w), but missing one sample at t8w (n=37), and stored at −80°C. Plasma samples were analyzed for 92 cardiovascular disease related protein biomarkers by Olink Proteomics (Olink Target 96 Cardiovascular III panel, Uppsala Sweden). The data received was QCed by excluding all proteins with a missing data frequency of >15% and samples with an Olink warning QC outcome from analysis. All samples with a low detection rate were given the limit of protein detection value for the specific protein. Using these QC parameters, none of the proteins had to be excluded.

### Relating clinical variables to cell type abundance, gene and protein expression

Cell type proportions and gene and protein expression profiles were associated with peak CK-MB values in STEMI patients. For gene expression associations we used MAST, a two-part generalized linear model on the log-transformed count matrix, and correcting for age, sex, 10X chemistry and 10X lane. For cell type proportion and protein expression associations, we first conducted a centered-log-ratio or log-transformation, respectively, after which linear regression was conducted, correcting for sex and age.

### pQTL analysis

For STEMI patients individual Olink plasma protein levels were linked to specific genetic variants (pQTL mapping) at each of the 3 timepoints (t0, t24h, t8w) using the eQTL mapping pipeline (https://github.com/molgenis/systemsgenetics/wiki/QTL-mapping-pipeline). We limited ourselves to the 241 variants that were previously found to show a genome-wide significant GWAS signal for coronary artery disease (CAD) (37). pQTLs were mapped per time point, only testing the SNPs within a 100kb window of the transcription start site of the gene transcribing the assessed protein. The significant pQTLs in STEMI patients in at least one timepoint were then assessed in controls by mapping these pQTLs in 1142 individuals from the general population cohort Lifelines Deep, in which the same Cardiovascular Olink panel was previously measured (38) and genotype data was available. Genetic variants were converted to Trityper format using GenotypeHarmonizer (https://github.com/molgenis/systemsgenetics). To assess whether the pQTL effect strength was influenced by the timepoint after the STEMI (t0, t24h, t8w), we then conducted an interaction analysis by fitting a generalized linear model correcting for age and sex as fixed effects, and the participant as a random effect.

### Statistical analysis

Discrete variables were represented as frequencies and percentages. Continuous variables with a normal distribution were summarized as mean ± standard deviation and if skewed were represented as median with interquartile range. Spearman correlation was used for non-normally distributed variables. Differences in protein levels were calculated using the Wilcoxon signed rank test. For DE gene and protein analysis, and their correlation to peak CK-MB values, multiple testing correction was performed using Holm-Bonferroni. We considered a corrected P-value <0.05 statistically significant. Statistical analysis and the figures were created using R Core Team (2020).

## Results

### Population characteristics of the STEMI patients and controls

To dissect the immune response during and after a MI, PBMCs and plasma from 38 patients with a first STEMI were collected at the time of hospital admission (t0), and 24h (t24h) and 6-8 weeks (t8w) after the event (**Figure 1**). The patient characteristics during hospital admission are described in **Table 1**. On average, STEMI patients were 60±11 years, had a median BMI of 26.4 (SD 3.5) kg/m^2^ and the majority were males (84%). Most patients presented with one stenosed vessel (55%), and in 39% of the patients the affected vessel was completely occluded (TIMI 0 flow). These 38 STEMI patients were compared to 38 age- and sex-balanced controls from the LifeLines DEEP cohort whose scRNA-seq data was previously published (19). These controls showed comparable age-(59.2 years) and sex-characteristics (84% male), but a slightly lower BMI (25.4 (SD 3.3) kg/m^2^) than the STEMI patients (**Table 1**).

### Single-cell profiling of immune cells in STEMI patients and controls

The collected PBMCs from the STEMI patients at three different timepoints post-STEMI (t0, t24h, t8w) and controls from the LifeLines cohort (22), were then used for 10X Genomics scRNA-seq analysis. A combination of 10X Genomics v2 and v3 chemistry reagents were used to capture an average of 842 cells per individual per condition (v2: 831 genes/cell, v3: 1,548 genes/cell) in STEMI patients, and an average of 891 cells per individual in controls (v2: 1,012 genes/cell, v3 1,931 genes/cell) (**Table S2**). Souporcell was used to demultiplex and to identify the doublets coming from different individuals (25). Souporcell clusters were correlated to individuals by correlating the cluster genotypes with donor genotypes using Spearman correlation. This revealed that by mistake we omitted loading cells for one donor at the t8w condition and that on average, 10.0% of the cells were assigned as doublet or inconclusive. Due to technical differences between v2 and v3 chemistry, determination of quality control (QC) thresholds and analyses were performed separately per chemistry. Low quality cells were excluded, leaving 129,873 cells (95,995 diseased and 33,878 control) in the final dataset used for analyses (see Methods). UMAP dimensionality reduction and KNN-clustering was then applied on the normalized, integrated count data, allowing the identification of cell types.

### Monocytes and NK cells show compositional changes after a STEMI and compared to controls

We identified 10 major cell types in the PBMC fraction of STEMI patients and controls, including the B, CD4^+^ T, CD8^+^ T, DC, HSPC, monocyte, NK, plasmablast, platelet and other T cells (**Figure 2a**). At higher resolution, these major cell types could be further splitted in 29 minor cell populations, including different subsets of T cells (regulatory T cells (Treg), CD4^+^ and CD8^+^ T-naive, CD4^+^ and CD8^+^ T central memory (TCM), CD4^+^ and CD8^+^ T effector memory (TEM), CD4^+^ cytotoxic (CTL), CD4^+^ and CD8^+^ proliferating, Mucosal associated invariant T cell (MAIT), double negative T (dnT), gamma-delta T (gdT), innate lymphoid cells (ILC), B cells (B naive, B intermediate, B memory), NK cells (NK dim, NK bright, NK Proliferating), monocytes (classical (cMono) and non-classical monocytes (ncMono)) and dendritic cells (AXL+ SIGLEC+ Dendritic Cell (ASDC), conventional DC1 (cDC1), cDC2, plasmacytoid DC (pDC)) (**Figure 2b**).

**Figure 2.**
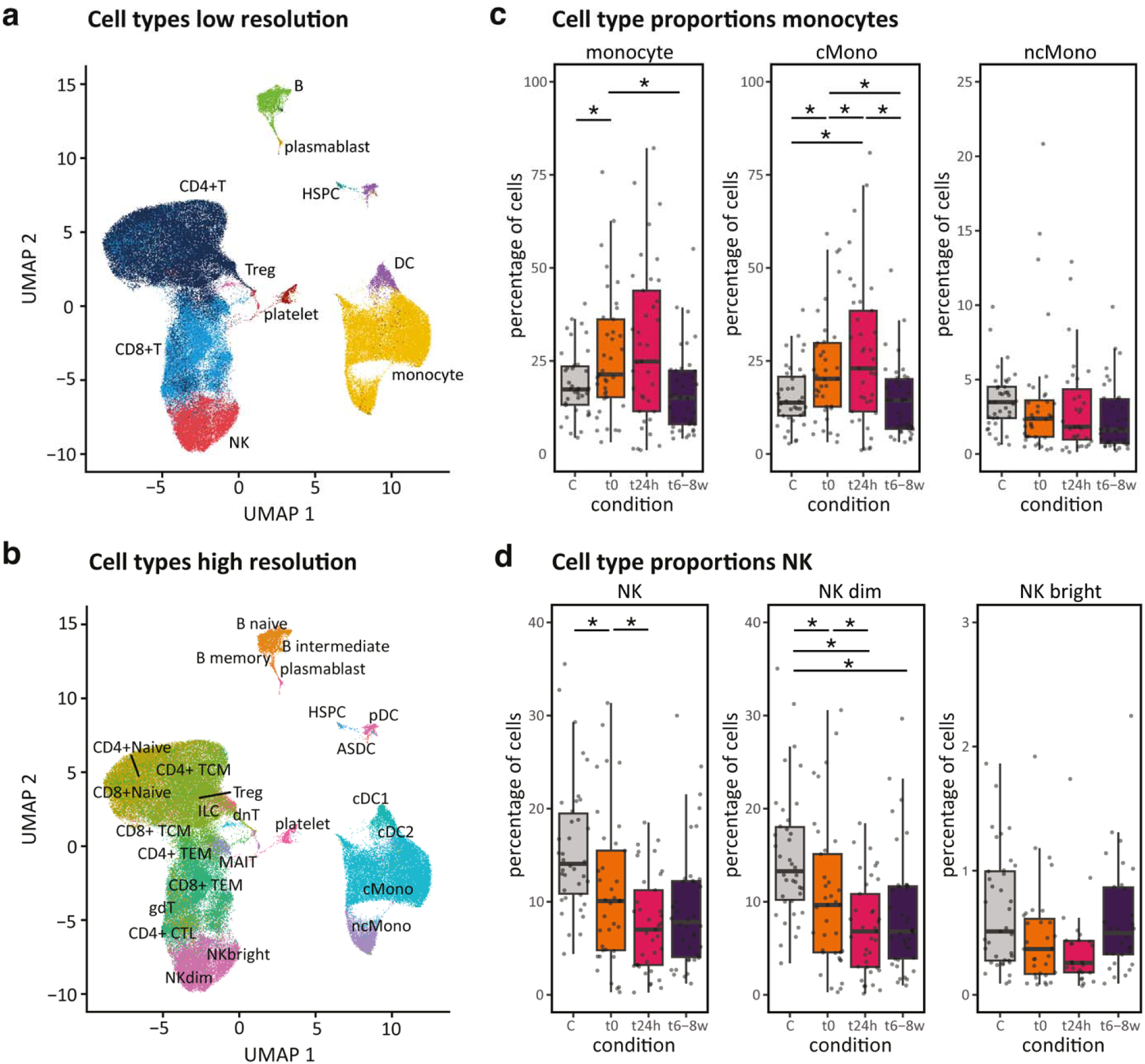
Cell type composition in STEMI patients over time and in controls. All PBMCs of STEMI patients at timepoints t0, t24h and t6-8w and controls are presented in UMAPs in figure **a** and **b**. **a**. showing 10 major cell clusters in the PBMC fraction **b**. showing 29 minor cell clusters **c**. Proportions of monocytes (major class and its two minor subtypes) and **d.** NK cells (major class and its two minor subtypes) in STEMI patients over time (0 hours - t0, 24 hours - t24h, and 6-8 weeks - t8w and in controls C), with significant changes in proportion, denoted at **\*** (P < 0.05). On higher cell type resolution, classical monocytes and NKdim cells appear to account for the change in proportion seen on lower resolution. cMono; classical monocytes, ncMono; non-classical monocytes, NK dim and NKbright.

To conduct a robust cell type composition analysis between the different conditions (STEMI patients at t0 compared to controls, STEMI patients over time), we initially focused our analyses on the 6 most abundant cell types (≥1% of the total PBMCs in each of the tested conditions): B cells, CD4^+^ T cells, CD8^+^ T cells, DCs, monocytes and NK cells (**Figure S4, Table S2)**. After which, we zoomed in at the higher resolution subcell type compositional changes to define the subtypes that contributed to these changes in composition (**Figure S5**).

Compared to controls, monocytes from STEMI patients during admission showed a relative increase (p=2.0×10^-5^) and NK cells a relative decrease in their abundance (p=9.6×10^-3^) (**Figure 2c, 2d, Table S3**). Zooming in further, we observed that these changes could be attributed to compositional changes of the cMono (p=2.0×10^-5^) and the NK dim (p=1.3×10^-2^) subcell types (**Figure 2c, 2d, Table S3**).

Similarly, in STEMI patients during the course of their disease (t0, t24h, t8w), we observed relative composition changes in the monocytes (decrease t0-t8w: p<5.0×10^-3^) and NK cells (decrease t0-t24h: p=9.0×10^-3^) (**Figure 2c, 2d**), but also in the CD4^+^ T cells (increase: t0-t8w: p=4.0×10^-3^) (**Figure S4**). These were again attributed due to changes in the cMonos (p=2.0×10^-5^) and NK dim cells (p=1.1×10^-2^), whereas none of the other minor cell types showed changes in composition during the disease course (**Figure S5**).

### Largest gene expression changes occur in monocytes and NK cells after a STEMI and in comparison to controls

To assess gene expression differences in STEMI patients at time of hospital admission versus controls, we performed DE gene analysis on the six major cell types using MAST (**Table S4**). Of all cell types, monocytes (846 DE genes), followed by NK cells (553 DE genes) showed the largest and B cells the least amount of DE genes (130 DE genes) (**Figure 3a**). The majority of the DE genes were uniquely identified in only one major cell type (**Figure 3b, 3e**). Looking at the monocytes, the cell type with most total and unique DE genes, we observed pathway enrichment among upregulated genes in STEMI patients involving various pro-inflammatory pathways, including IFN signaling, interleukins (IL-1, IL-6) and all toll-like receptors (**Table S5**).

**Figure 3.**
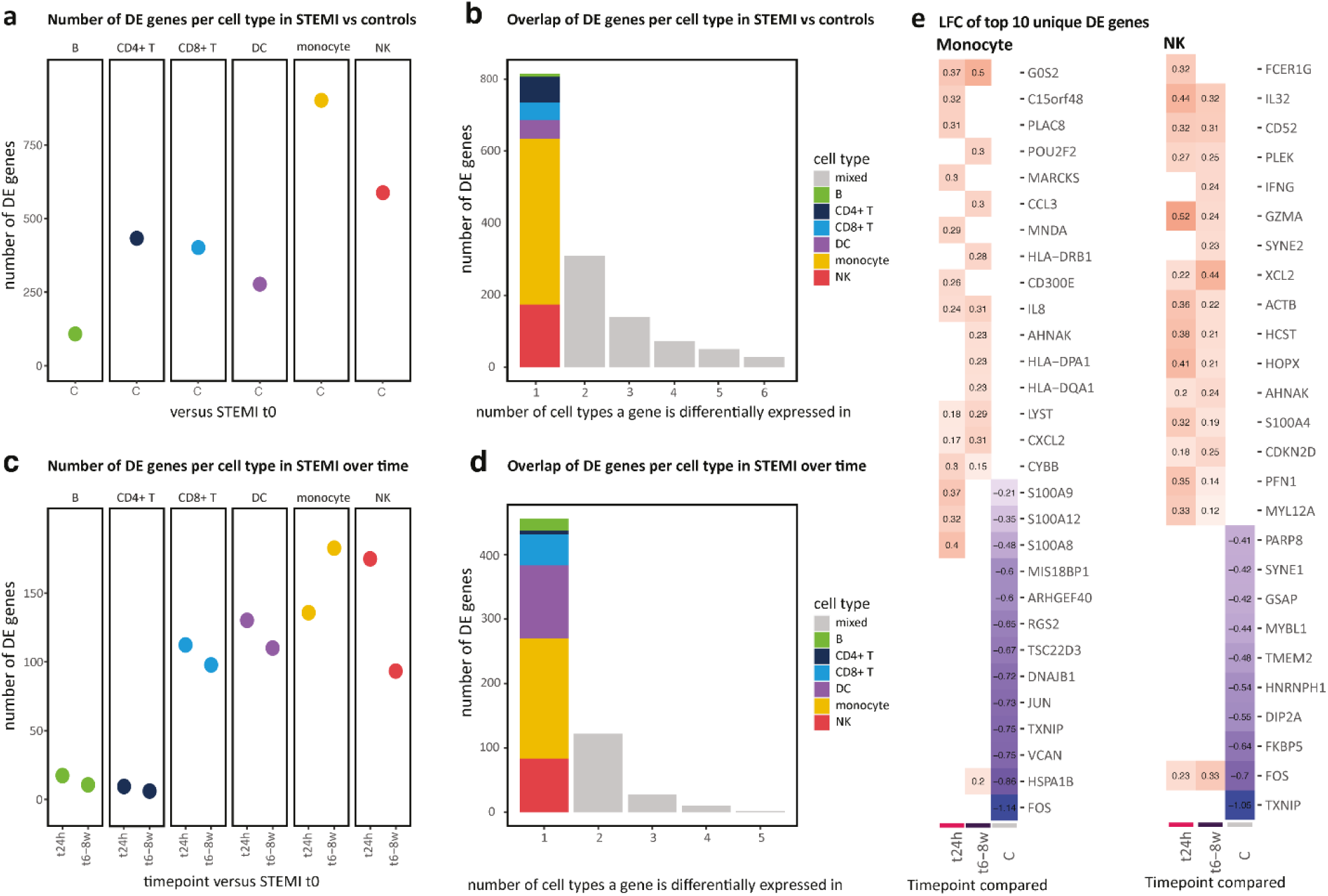
Differential expressed genes in STEMI patients and in controls. **a**. The number of DE genes per cell type in STEMI patients at hospital admission (t0) compared to controls (C) or **c.** in STEMI patients over time. **b**. Bar plot showing the overlap of DE genes in cell types at t0 compared to controls or **d**. in STEMI patients over time (taking all the DE genes that were significant in at least one of the comparisons (t0-t24h or t0-t8w)). The number of participants in each condition is 37, 37, 38 and 38 for t0, t24h, t8w and C respectively. **e.** Heatmap showing the log fold change expression of significant unique DE genes (1^st^ column in **b** and **d**) in monocytes (left) and NK cells (right). All comparisons are against t0, showing the top 10 upregulated genes in t24h and t8w, and the top 10 downregulated genes in C. LFC is only given when a significant difference with t0 was found in that condition.

Subsequent comparison of the gene expression levels of STEMI patients at hospital admission compared to later time points revealed the largest amount of DE genes in NK cells and in monocytes, respectively 24h and 6-8 weeks after STEMI (**Figure 3c, Table S4**). Specifically looking at the pathways underlying the upregulated genes at these cell type-timepoint combinations, we observed that many of the NK DE genes at t24h were involved in interactions with platelets (activation, degranulation, response to platelet cytosolic Ca2+), whereas many of the monocyte DE genes at t8w were involved in pro-inflammatory signaling (IFN-gamma, IL-1) (**Table S5**). Remarkably, all major types showed a decrease in DE genes going from 24h to 6-8 weeks after STEMI, except for the monocytes (**Figure 3c**). And as before in comparison to controls (**Figure 3b**), monocytes also showed the largest proportion of unique DE genes over the course of the disease (**Figure 3d, 3e**). Together with, but independently of, the observed cell type composition changes (**Figure 2c, 2d**), these results indicate that both the monocytes and NK cells seem to play a pivotal role in the immunological changes that are occurring upon a STEMI.

### CD8+ T and NK cells show most potential differential cell-to-cell communication during the acute, and monocytes during the chronic phase after STEMI

Next, we defined how cell-to-cell communication interactions among immune cell types could underlie these observed gene expression changes in STEMI patients over time. For this, we used NicheNet, a cell-to-cell communication tool that identifies potential cell-cell interactions by assuming that ligand-receptor interactions can be predicted based on the expression of the ligand in one cell type and downstream gene expression changes of a known ligand-receptor interaction in another cell type (**Table S6**). This analysis revealed that in the acute phase of the disease (t0-t24h) in- and outcoming communication was balanced among the cell types (**Figure 4a**). In contrast, in the chronic phase of the disease (t24h-t8w) the T cells and DCs were mostly sending communication, while monocytes and B cells were mostly receiving it (**Figure 4b**). Although from the comparable number of incoming interactions for a specific cell type it may seem that the involved ligands are mostly shared among cell types, most of these ligands were actually uniquely involved in the communication by just one cell type (**Figure 4c, 4d**). Looking at the number of ligand-receptor interactions that were associated with changes in downstream signaling, we observed both NK and CD8^+^ T cells to be the largest communicators during the acute phase (**Figure 4a**), whereas monocytes were the main communicators during the chronic phase of the disease (**Figure 4b**). These results align well with our DE analyses, which also indicated the largest impact on expression changes of STEMI on NK cells early and on monocytes later in the disease.

**Figure 4.**
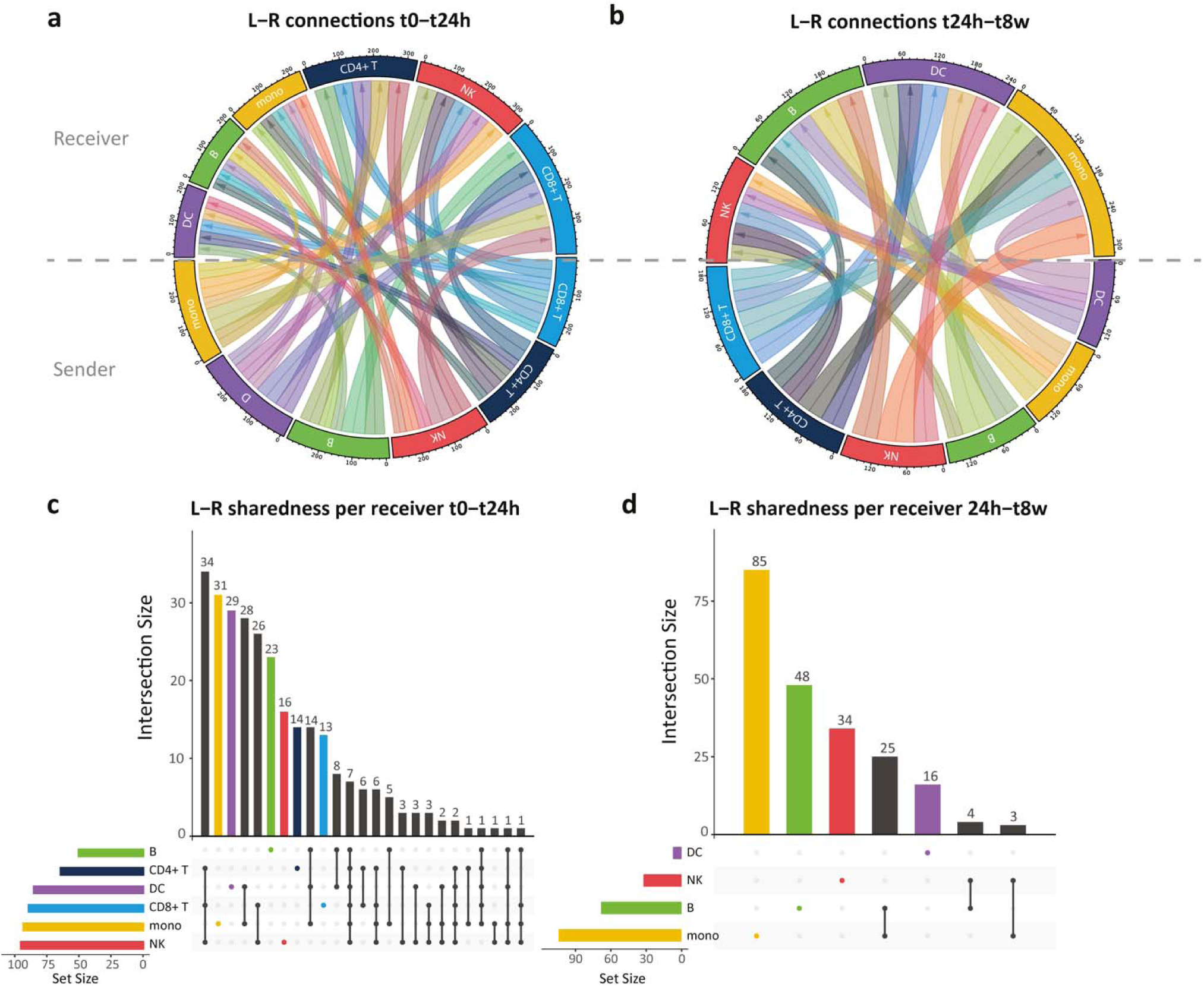
Differential putative cell-to-cell communication after STEMI. Circos plots depicting the differential incoming (receiver) and outgoing (sender) putative cell-to-cell communication at the t0-t24h (**a**) or t24h-t8w (**b**) period. An active cell-to-cell communication link is counted as being a ligand-receptor (LR) link that has resulted in differential downstream gene expression. For each active LR link in **a** (t0-t24h) and **b** (t24h-t8w), the sharedness of ligands among the major cell types is depicted in **c** and **d**, respectively. The number of participants in each condition is 13, 13, 14 and 7 for t0, t24h, t8w and C respectively (V3 chemistry data).

### Proteins known to be involved in cardiovascular disease show changes over time after STEMI

While our previous analyses at the mRNA level help us to disentangle the molecular processes that immune cells undergo after STEMI, changes in plasma protein levels may provide us additional insights on the systemic consequences of a STEMI and can be more easily monitored in clinical follow-up of patients. Therefore, next, using the Olink Target 96 Cardiovascular III panel, we analyzed in each of the three timepoints (t0, t24h, t8w) within STEMI patients the plasma levels of 92 proteins that are known or exploratory human cardiovascular and inflammatory markers.

Targeted differential protein analysis revealed that 14 out of 92 assessed plasma protein levels changed during the first 24h (t0-t24h: 8 up, 6 down) and 28 out of 92 during the first 8 weeks (t0-t8w: 22 up, 6 down) after STEMI. During the first 24h, the top three upregulated proteins were NT-proBNP (mean LFC= 0.74, p=2.1×10^-7^), IL1RL1 (mean LFC=0.18, p=7.3×10^-4^), and CHI3L1 (mean LFC=0.16, p=6.7×10^-4^) (**Figure 5**, list of all DE proteins shown in **Table S7**). Both upregulation of NT-proBNP and the soluble form of IL1RL1, a member of the IL-1 receptor family, are well known proteins that are upregulated in response due to increased wall stress during STEMI (39). Moreover, both are independent predictors of heart failure and cardiovascular death (40,41). CHI3L1, also known as YKL-40, is an extracellular matrix protein which is involved in several stages of CAD. It induces maturation of monocytes to macrophages in the early phase of atherosclerosis, but also promotes atherosclerotic progression and complications like plaque rupture (42). So, the upregulation of this protein might be due to the cause of MI (i.e. plaque rupture), whereas NT-proBNP and IL1RL1 reflect the mechanical stress to the cardiomyocytes due to MI.

**Figure 5.**
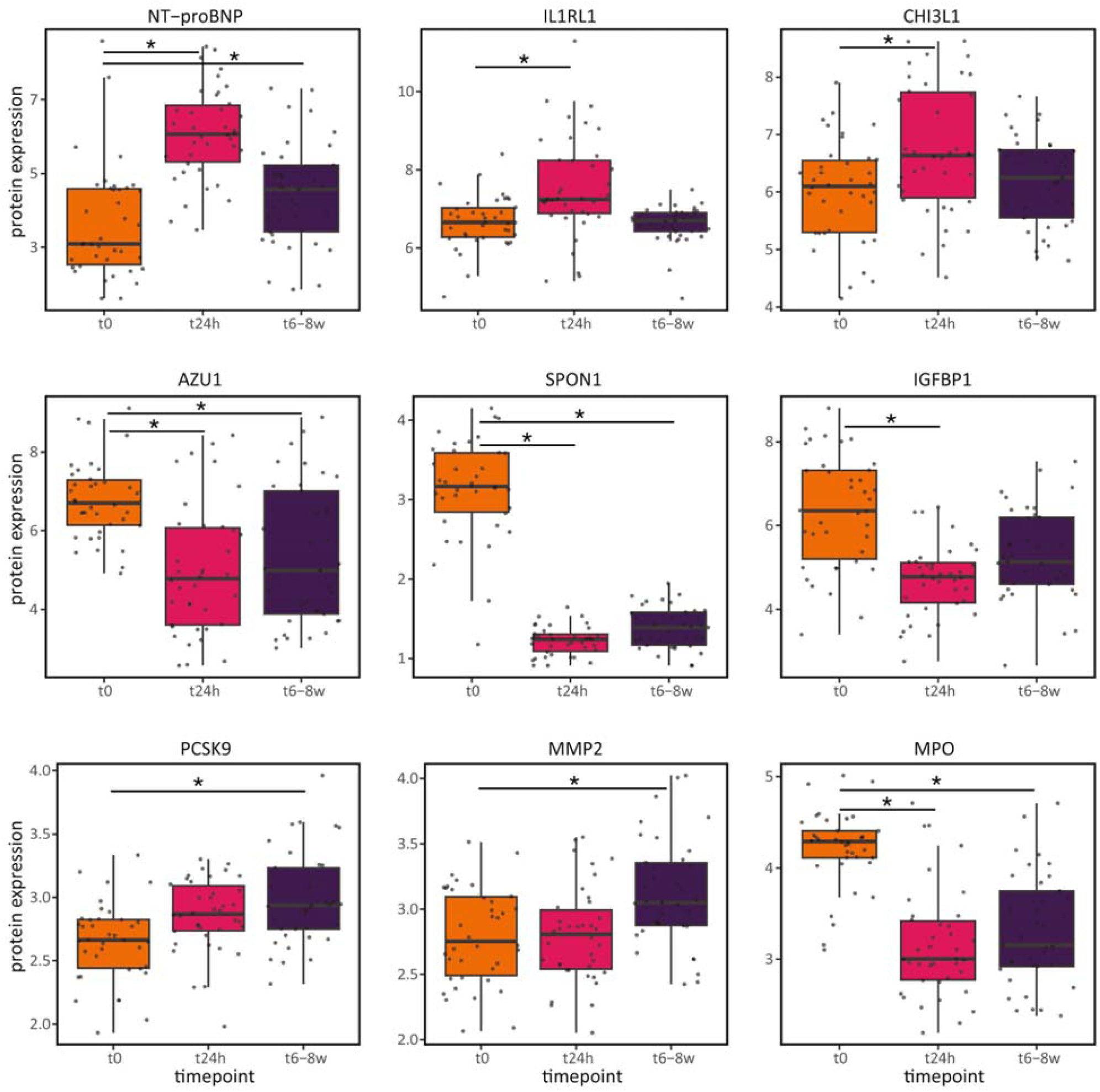
Differentially expressed proteins after STEMI. Top three highest up- and downregulated proteins at 24h (t24h) and 6-8 weeks (t8w) after STEMI compared to time of hospital admission (t0). Significant changes in protein level after Bonferroni multiple testing correction are denoted by * (P<0.05). The number of participants in each condition is 38, 38 and 37 for t0, t24h and t8w respectively.

The top three downregulated proteins during the acute phase were SPON1 (mean LFC=-1.4, p=6.7×10^-10^), AZU1 (mean LFC=-0.44, p=3.3×10^-6^) and IGFBP1 (mean LFC=-0.43, p=9.9×10^-6^). Each of these proteins have been previously associated with processes preceding a STEMI (e.g. atherosclerosis for AZU1 (40) and IGFBP1 (44,45)) or that are the consequence of unsuccessful STEMI treatment (e.g. worsened systolic heart function for SPON1 (46). Therefore, we suspect their downregulation during the acute and chronic phase may be an indication of effective treatment by PCI after STEMI, resulting in restoration of blood flow and eventually preservation of heart function.

During the chronic phase, the top three upregulated proteins were NT-proBNP (LFC= 0.3, p= 1.3×10^-2^), PCSK9 (mean LFC=0.18, p=6.0×10^-2^) and MMP2 (mean LFC=0.16, p=2.6×10^-3^). The increased NT-proBNP levels may reflect the further increased wall stress in the chronic phase after STEMI. PCSK9 plays an important role in degradation of the LDL-receptor (LDLR) (47) and the observed chronic increase of PCSK9 might therefore reflect an ongoing state of hyperlipidemia. MMP2 is a matrix metalloproteinase-2 that acts in the fibrotic pathway (48), and is involved in cardiac remodeling post STEMI (49).

The top three downregulated proteins during the chronic phase were SPON1 (mean LFC=-1.2, p=1.3×10^-9^), MPO (mean LFC=-0.36, p=2.0×10^-6^) and AZU1 (mean LFC=-0.35, p=5.6×10^-4^). Similar to the acute phase, both SPON1 and AZU1 continued to be downregulated. Additionally, MPO was found downregulated, a protein involved in the acute innate inflammatory response which contributes to plaque destabilization through local oxidative tissue injury (50,51). Together, these changes seem to indicate the long-term restoration of heart function and blood flow by PCI.

In a very comparable cohort of 48 STEMI patients, using the same Olink CVDIII panel, Kulasingam et al. compared the plasma proteins at hospital admission with those 3 months after STEMI. Interestingly, they found 29/92 proteins to be differentially expressed during this timeframe (52). Out of these 29, 12 were also found in our study, and all with the same direction of effect, indicating the robustness of our results (Table S7). These replicating differential proteins were mainly involved in well-known processes that occur after STEMI, such as the immune and inflammatory response, cell adhesion and tissue remodelling.

### Gene and protein expression profiles were not associated to biochemical-defined infarct size

STEMI patients can already widely differ from each other when they are presented with symptoms in the clinic. While the patient inclusion and exclusion criteria of our study (**Methods**) reduced the amount of clinical variation in our study (**Table 1**), many aspects remain that could underlie the observed donor-to-donor variation in the measured molecular phenotypes, including age, sex, infarct size and genetics. To assess whether variation in infarct size may correlate with the individual molecular data layers, we used plasma peak CK-MB levels as proxy. In clinical practice, plasma CK-MB levels are measured regularly after STEMI to monitor myocardial damage. This biomarker reaches a peak within 24 hours, and the peak value is an estimation of the infarct size and predicts left ventricle dysfunction (53). First, we used plasma peak CK-MB levels to assess whether PBMC cell type proportions and monocyte gene expression levels were associated with biochemical infarct size. We focussed on gene expression of the monocytes as this was the cell type with the largest amount of total and unique DE genes. However, neither cell type proportions, nor monocyte gene expression levels showed a significant association with peak CK-MB at any of the tested timepoints (t0, t24h or t8w). Next, we assessed whether peak CK-MB values were associated with inflammatory plasma protein levels in the acute and chronic phase after STEMI using simple linear regression. Of all 92 proteins, only NT-proBNP was positively associated with peak CK-MB in the acute phase (t24h: r^2^ adjusted=0.49, p=2.8×10^-3^) (**Table S8**). NT-proBNP, a marker for myocardial wall stress, is commonly measured in clinical practice to assess left ventricular dysfunction (54,55). As there is a direct relationship between the level of myocardial damage and the amount of left ventricular dysfunction, NT-proBNP and CK-MB levels are expected to be correlated during the time CK-MB peaks (∼24h after STEMI) (39,56).

### Genetic risk variants for CAD show disease- and condition-dependent effects on plasma protein levels of known drug targets

Beyond infarct size, genetic variation among patients, can be an important contributor to the observed differences in molecular response among patients. Therefore, it is important to take genetic variation into account when assessing a patient’s disease course and defining patient-tailored treatment. As the measured plasma protein levels provide a pool of potential direct therapeutic targets, we assessed the effect of genetic variation on these. To provide a direct clinical link, we specifically focussed on those genetic variants that were previously found to be associated with CAD risk (37). For three out of 92 plasma proteins, we detected a significant pQTL in the STEMI patients in at least one of the three timepoints (t0, t24h, t8w) (**Table S9**). For these three pQTLs, we assessed their disease-specificity by comparing pQTL effect sizes with those in a control cohort of 1142 individuals from the general population. This revealed one pQTL effect that was significant in all conditions and whose effect size was comparable among both patients (independent of time point) and controls: SNP rs6686750 affecting IL6R plasma protein levels (STEMI patients: genotype β=0.42, p=1.6×10^-5^, **Figure 6a**). The two other pQTLs were disease- or even condition-specific: SNP rs8124182 affecting MMP9 plasma protein levels was found specifically in STEMI patients (STEMI patients: genotype β=-0.68, p=0.016,**Figure 6b**) and SNP rs10422256 affecting LDLR plasma protein levels was found only in STEMI patients at t24h (STEMI patients: genotype x t24h interaction β=-0.29, p=0.022, **Figure 6c**). These results indicate the importance of taking genotype into consideration, when studying the relationship between STEMI and molecular phenotype.

**Figure 6.**
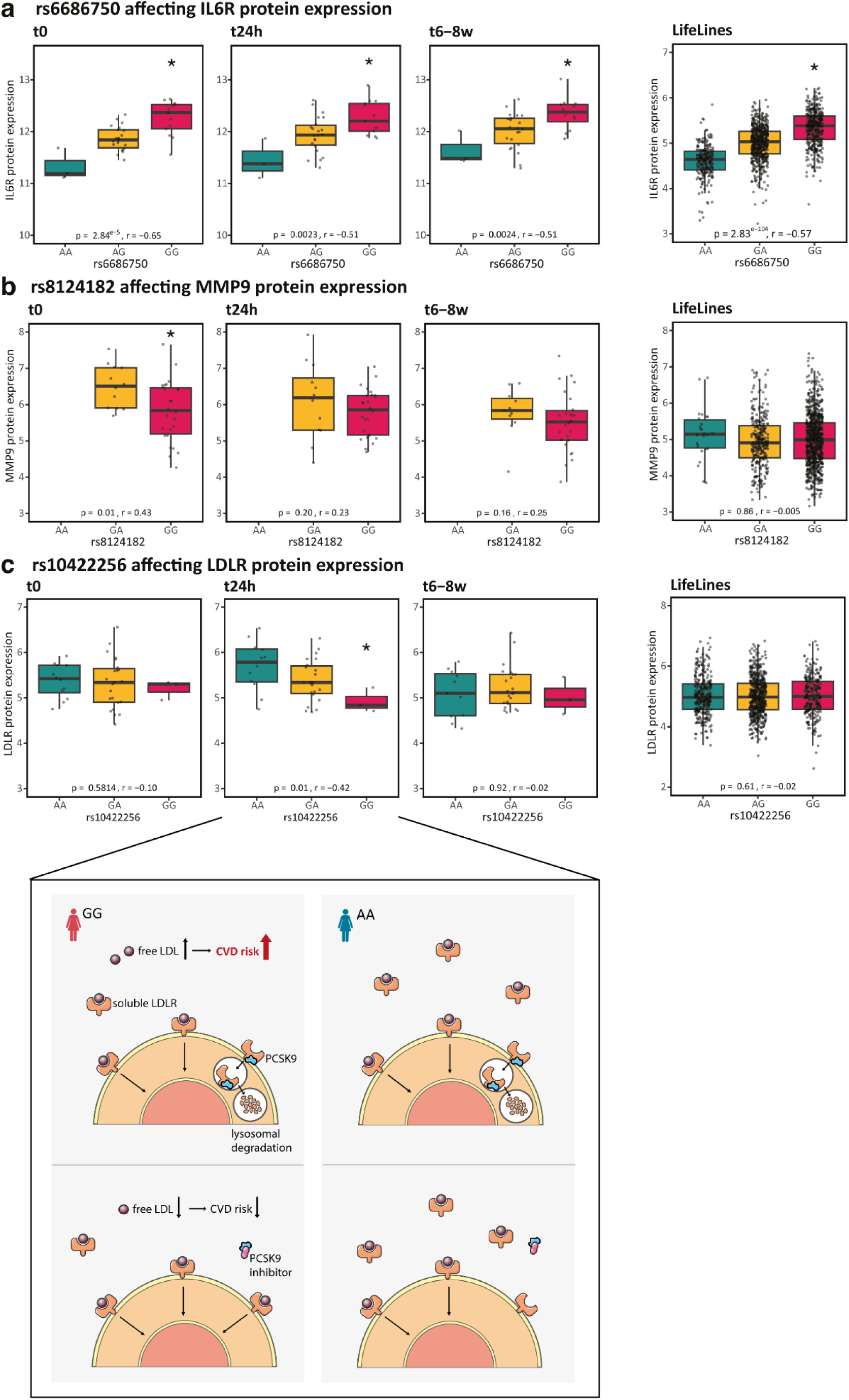
Plasma protein Quantitative Trait Loci (pQTL) in STEMI patients compared to controls (LifeLines) show both condition-dependent and condition-independent changes over time. Plasma pQTLs were measured in 38 STEMI patients at time of hospital admission (t0), 24 hours (t24h) or 6-8 weeks (t8w) after STEMI and compared to 1142 controls from the general population cohort Lifelines Deep (18). Three out of 92 plasma proteins showed a pQTL effect that *a.* was significant in all conditions (IL6R), *b.* specific for STEMI patients (MMP9) or *c.* even specific for one timepoint in the STEMI patients (t24h: IL6R). The number of samples for each genotype group can be found in *Table S10*.

## Discussion and Conclusion

In this study, we have provided an unbiased longitudinal overview of the single-cell immune response in STEMI patients and compared this to age- and sex-balanced controls. We first mapped changes in the cell type composition and gene and plasma protein expression level. We then determined the overarching pathways that were enriched and the cell types being involved in these changes as a result of cell-to-cell communication. Thereafter, we determined whether these changes were correlated with an individual’s (biochemically-measured) infarct size, genotype or disease phase. Together, all these analyses indicated an important role for the classical monocytes and NKdim cells during STEMI, both in terms of cell type compositional and gene expression changes.

Our observation that monocytes play a pivotal role in STEMI is in line with a previous scRNA-seq study of 10 STEMI patients (with and without plaque rupture) that were analyzed at time of hospital admission (57). While this study did not observe any differences in cell type composition between STEMI patients with or without plaque rupture, they did observe that most cell-cell communication occurred from and to monocytes. Our study now adds additional insights, because of the longitudinal character, the added layer of protein information, the increased sample size, and the availability of controls to compare with. The recently published spatial multi-omic map of human MI, in which cardiac structure and specific cardiac cells were studied in spatial context, underlined the role of immune cells in cardiac repair following STEMI (17). While that study used cardiac tissue, and we studied the response of circulating immune cells, it remains clear that gaining more insights into the immune response after STEMI is key for future drug therapies.

One such insight relevant for future drug therapies that our study has now provided, is in relation to the use of IL-1⍰ blockers in MI. Our study showed chronic upregulation of the IL-1 signaling pathway specifically in the monocytes of STEMI patients. The CANTOS trial showed that IL-1⍰ inhibition with canakinumab led to lower cardiovascular event rates, but also higher fatal infections (4). Our insights into the IL-1 pathway could aid in targeting specifically monocytes, and thereby potentially preventing the occurrence of serious side effects. Furthermore, a study in IL-1R knock-out mice showed that these mice, which cannot respond to IL-1, were protected from hypertrophy, fibrosis, and diastolic dysfunction (58). This may indicate that the observed chronic upregulation of the IL-1 signaling pathway after STEMI reflects an already ongoing process of developing heart failure and eludes the potential to broaden the use of IL-1 inhibition for both atherosclerotic events and fibrotic processes.

Additionally, our study indicated the importance of considering both genetic variation and disease phase when assessing the molecular consequences of a STEMI, as several of the protein expression levels were affected by a combination of these parameters. Importantly, by combining molecular layers, our integrative analysis revealed insights in several therapeutic targets for which drugs are already available and are currently tested in clinical trials.

The first example for a promising drug target in the context of CVD, involves targeting IL-6 signaling. IL-6 signaling can be activated through three different modes of signaling, each having their own consequences: classical signaling, trans-signaling and trans-presentation (59). The anti-inflammatory classical signaling is activated by membrane-bound IL6R and gp130 (also known as IL6ST) within the same cell, the pro-inflammatory trans-signaling is regulated by soluble IL-6R and membrane-bound gp130, and the Th17-cell promoting trans-presentation signaling is induced by membrane-bound IL-6R (in DCs) and gp130 (in Th17 cells). Through cell-cell communication analyses with NicheNet (**Table S7**), we found that both the classical and trans-presentation pathways were differentially active during the first 24h of STEMI. While our pQTL analysis indicated that the trans-signaling pathway was affected by SNP rs6689206 regulating soluble IL6R levels both in STEMI patients as well as in controls (**Figure 6a**). Each of these IL-6 pathways can be targeted using different drugs (59), and based on our findings it seems that these drugs may be most beneficial during specific stages of the disease (first 24h after STEMI: for classical and trans-presentation IL-6 signaling, using drugs like anti-IL-6 (59,60) or anti-IL-6R, or in donors with a specific genetic background (patients with the GG genotype at SNP rs6689206 have the highest expression of IL-6R and thereby will naturally activate IL-6 trans-signaling the most, and as such are expected to benefit most of treatments targeting specifically this pathway, e.g. Olamkicept, soluble gp130).

Another interesting drug target for which we now provide additional insights is the LDLR. LDLR plasma levels act as scavengers for plasma LDL and can be indirectly targeted by PCSK9 inhibitors, which inhibit the degradation of membrane LDLR (47). As such, they are widely used to decrease LDL cholesterol and reduce the risk of cardiovascular events (47). Our DE protein analyses revealed that PCSK9 levels increased after STEMI (**Figure 5**), while our pQTL analysis revealed that 24h after STEMI individuals with the GG genotype (at SNP rs10422256) have a lower plasma protein level of LDLR independent of their PCSK9 levels (**Figure 6c**) and may therefore have higher free plasma LDL leading to increased cardiovascular disease risk (**Figure 6c**). Together, this indicates that individuals with the GG genotype at SNP rs10422256 may benefit the most from PCSK9 inhibitor treatment, especially in the first 24 hours of the disease. We expect that implementing such genetic and phase of the disease information will be important to maximize benefit of such patient-tailored therapies in future clinical practice.

Unfortunately, when focussing on how each of the molecular layers could be contributing to or being the consequence of biochemically-measured infarct size, we could not identify any direct relationship except for plasma NT-proBNP levels (**Table S9**) – a known marker for heart failure (54,55). We suspect this lack of association might have been the result of assessing each molecular parameter separately or not considering all potentially relevant clinical and donor variables (age, sex, smoking status, genetics, etc.) into the model (61). To enable such a model considering all these parameters, a larger sample size would be required. We expect such approaches to be feasible in the near future by performing meta-analyses across multiple (disease) population-based single-cell datasets that are uniformly processed in efforts such as those of the single-cell eQTLGen consortium (10).

In conclusion, this study highlights the importance of studying STEMI at cell-type-specific resolution, while taking genetic variation, disease status and phase of the disease into consideration. We expect that such an integrative approach as used here, will help to better grasp the molecular processes underlying STEMI and will be essential for the development of effective future therapies with reduced side-effects.

## Abbreviations

AZU1: Azurocidin
CAD: Coronary artery disease
CHI3L1: Chitinase-3-like-1
CITE-seq: Cellular indexing of transcriptomes and epitopes by sequencing
CK: Creatine kinase
CK-MB: Creatine kinase, myocardial band
cMono: Classical monocyte
EDTA: Ethylenediaminetetraacetic acid
HbA1c: Glycated hemoglobin
C: Control
IGFBP1: Insulin like growth factor binding protein
1 IL-1: Interleukin 1
IL6R: Interleukin 6 receptor
IL1RL1: Interleukin 1 receptor-like 1
LDL: Low-density lipoprotein
LFC: Log fold change
MAF: Minor allele frequency
MAST: Model-based analysis of single cell transcriptomics
MCV: Mean corpuscular volume
METC: Medical ethical review committee
MI: Acute myocardial infarction
MMP2: Matrix metalloproteinase-2
MPO: Myeloperoxidase
ncMono: non-classical monocyte
NT-proBNP: N-terminal pro-brain natriuretic peptide
PBMC: Peripheral blood mononuclear cell
PCI: Percutaneous coronary intervention
PCSK9: Proprotein convertase subtilisin/kexin type 9
pQTL: Protein quantitative trait locus
QC: Quality check
scATAC-seq: Single-cell assay for transposase-accessible chromatin sequencing
SNP: Single-nucleotide polymorphism
scRNA-seq: Single-cell RNA-sequencing
snRNA-seq: Single-nucleus RNA-sequencing
SPON1: Spondin-1
STEMI: ST-elevated myocardial infarction
TIMI: Thrombosis in myocardial infarction
UMAP: Uniform manifold approximation and projection
UMCG: University medical center groningen
UMI: Unique molecular identifier
VD: Vessel disease

## Acknowledgements

The images are created using Servier Medical Art and we are thankful to www.smart.servier.com for providing free online images.

## Data availability

Processed (de-anonymized) scRNA-seq data, including a text file that links each cell barcode to its respective individual, has been deposited at the European Genome-Phenome Archive (EGA), which is hosted by the EBI and the CRG, under accession number EGAD00001010064.

## Code availability

The code for Seurat v4 (https://github.com/satijalab/seurat), Eagle v2.x

(https://github.com/poruloh/Eagle), Souporcell v1.x (https://github.com/wheaton5/souporcell) and our in-house eQTL pipeline2 v1.4.0 (https://github.com/molgenis/systemsgenetics/tree/master/eqtl-mapping-pipeline) can be found at GitHub.

All custom code is also available via GitHub (https://github.com/molgenis/STEMI-scRNA-seq).

## Supplementary figures

**Figure S1.**
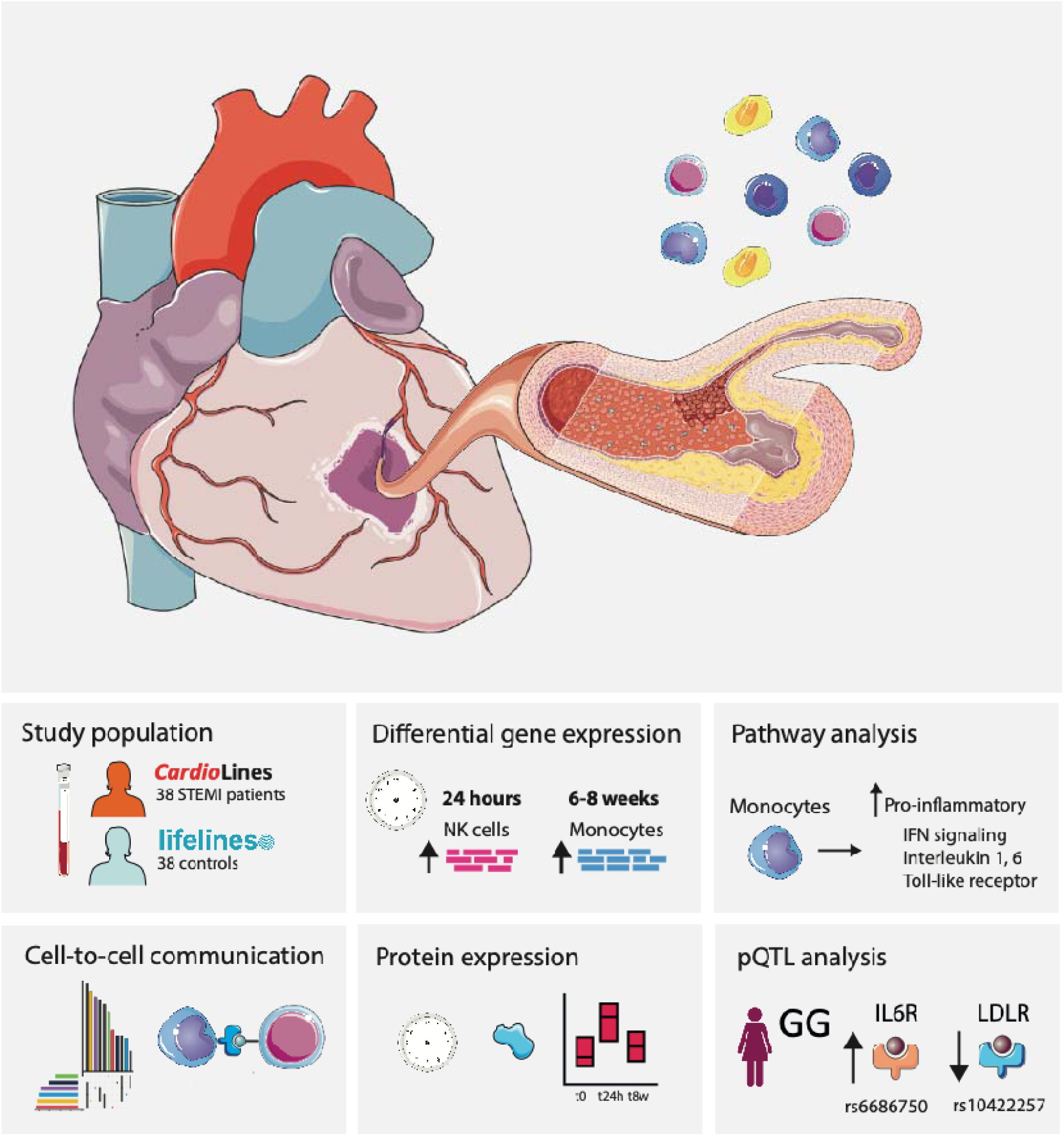
Overview of the study. Describing the cohort and the results per section.

**Figure S2.**
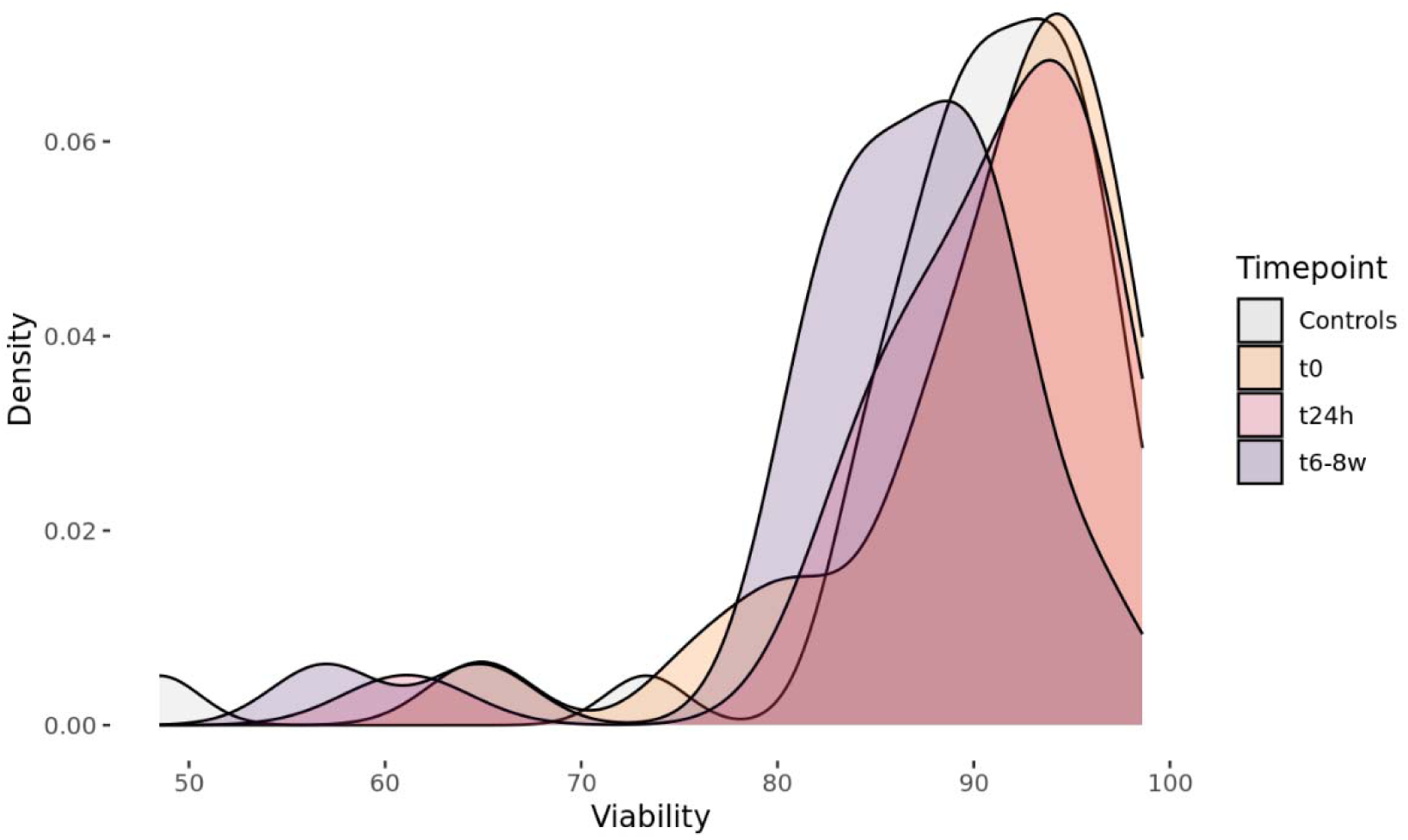
Histogram showing the PBMC cell viability as measured by Trypan Blue in STEMI patients over time (t0, t24h, t6-8w) and controls for cells prior to loading for scRNA-seq.

**Figure S3.**
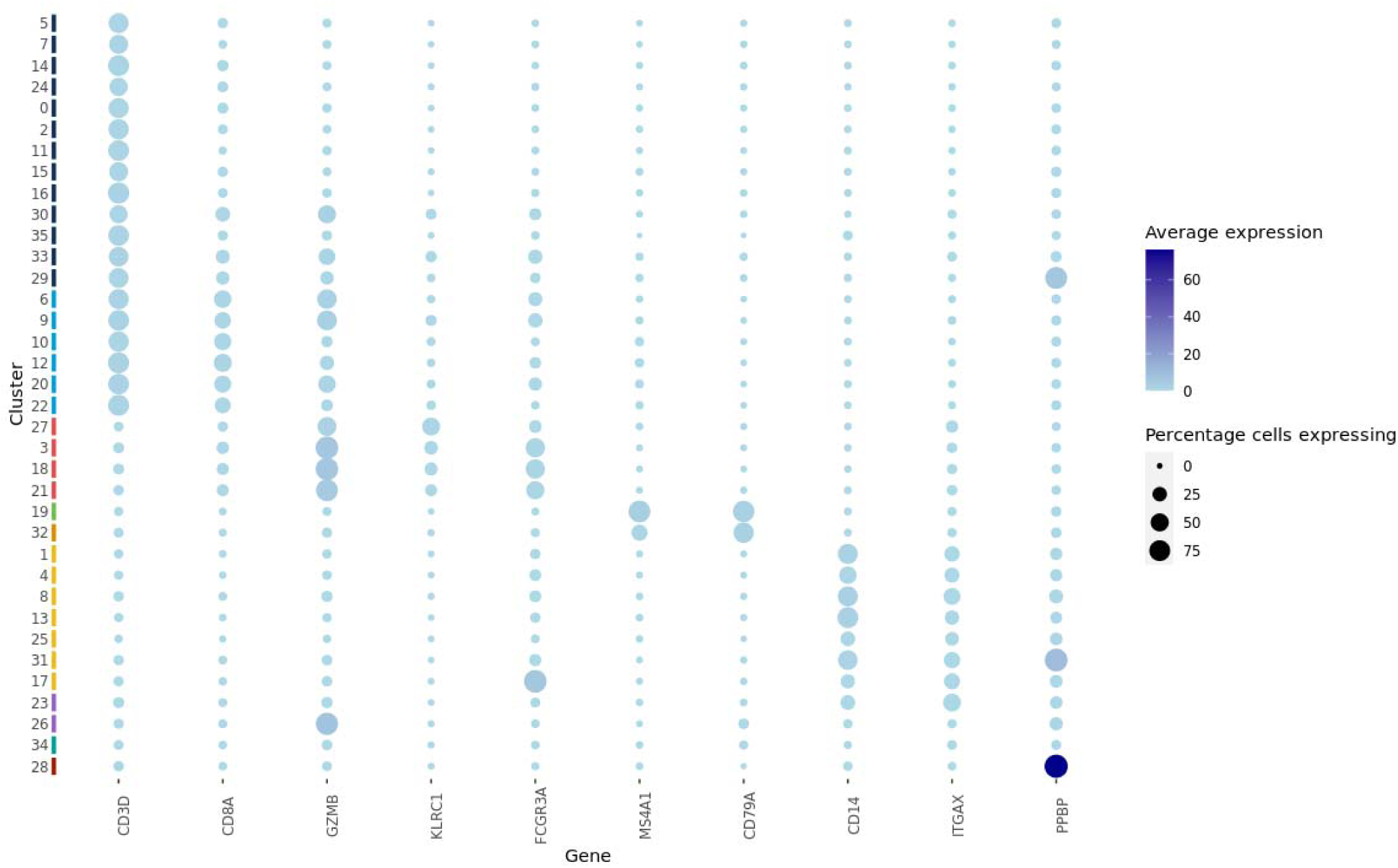
Dotplot showing marker gene expression per cell type. Each cluster was assigned the most common Azimuth-annotated cell type. Correct assignment was validated using marker gene expression (for each cluster, the dot color represents the average expression and the dot size the percentage of cells showing expression), and if needed clusters were re-assigned.

**Figure S4.**
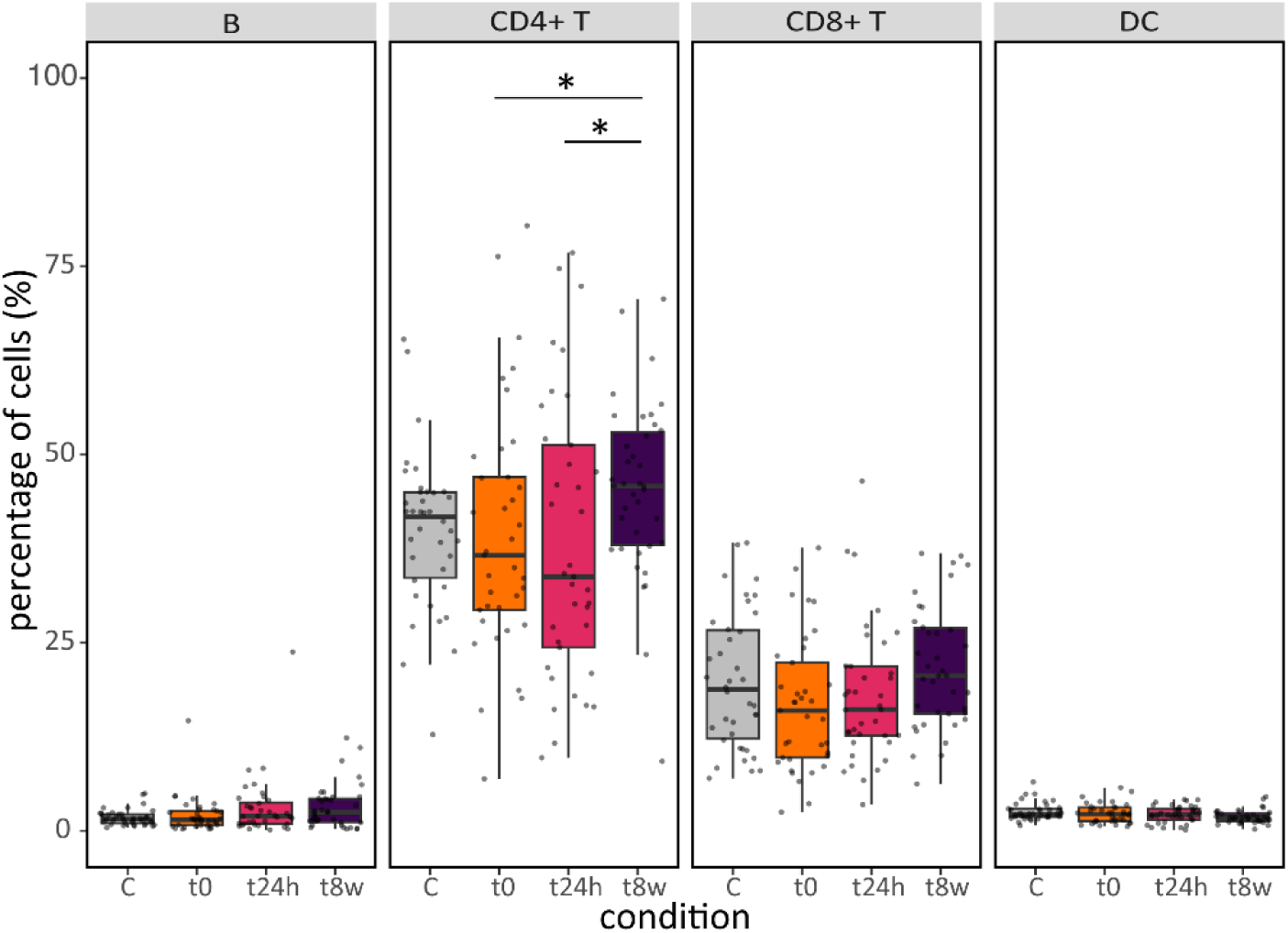
Proportions of major cell types in STEMI patients compared to controls (Cs) and over time at three different time points (t=0, t=24 hours and t=6-8 weeks post STEMI). Significant differences are denoted as **\***P < 0.05. n is 37, 37, 38 and 38 for t0, t24h, t8w and C respectively.

**Figure S5.**
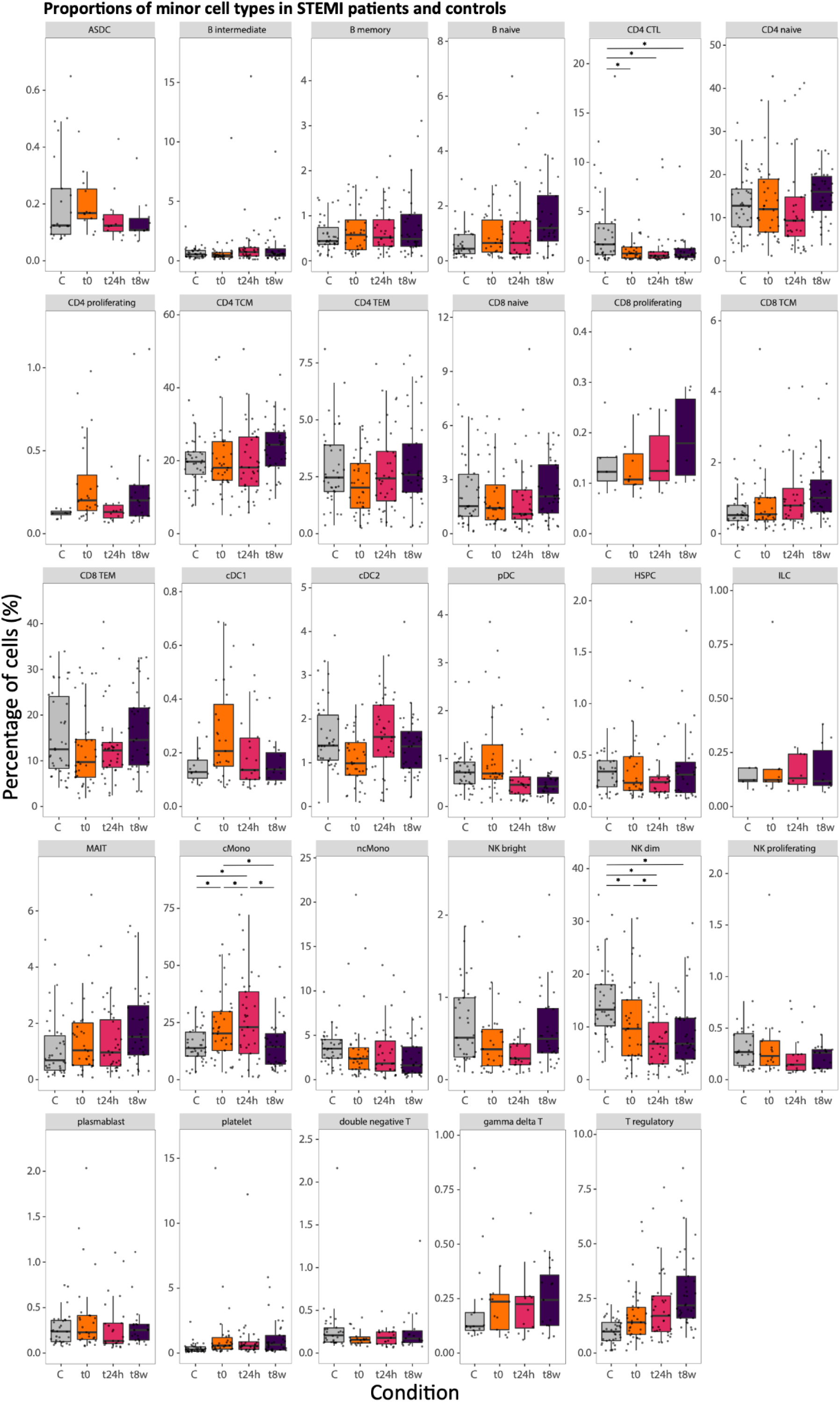
Proportions of minor cell types in STEMI patients compared to controls (C) and over time at three different time points (t=0, t=24 hours and t=6-8 weeks). ASDC; Axlp Sigle6p Dendritic Cell, CTL; Cytotoxic cell, TCM; T-central memory, TEM; T-effector memory, cDC; conventional dendritic cell, pDC; plasmacytoid dendritic cell, HSPC; Hematopoietic stem and progenitor cell, ILC; Innate lymphoid cell, MAIT; Mucosal associated invariant T cell, cMono; classical monocytes, ncMono; non-classical monocytes. Significant differences are denoted as **\***P < 0.05. n is 37, 37, 38 and 38 for t0, t24h, t8w and C respectively.

